# A translational multiplex serology approach to profile the prevalence of anti-SARS-CoV-2 antibodies in home-sampled blood

**DOI:** 10.1101/2020.07.01.20143966

**Authors:** Niclas Roxhed, Annika Bendes, Matilda Dale, Cecilia Mattsson, Leo Hanke, Tea Dodig-Crnkovic, Murray Christian, Birthe Meineke, Simon Elsässer, Juni Andréll, Sebastian Havervall, Charlotte Thålin, Carina Eklund, Joakim Dillner, Olof Beck, Cecilia E Thomas, Gerald McInerney, Mun-Gwan Hong, Ben Murrell, Claudia Fredolini, Jochen M Schwenk

**Affiliations:** Micro and Nanosystems, School of Electrical Engineering and Computer Science, KTH Royal Institute of Technology Stockholm, 100 44 Stockholm, Sweden; Science for Life Laboratory, School of Engineering Sciences in Chemistry, Biotechnology and Health, KTH Royal Institute of Technology, 171 65 Solna, Sweden; Department of Microbiology, Tumor and Cell Biology, Karolinska Institutet, 171 65 Solna, Sweden; Science for Life Laboratory, Karolinska Institutet, Department of Medical Biochemistry and Biophysics, Division of Genome Biology, Solna, 171 65, Sweden; Ming Wai Lau Centre for Reparative Medicine, Stockholm node, Karolinska Institutet, Solna, 171 65, Sweden; Science for Life Laboratory, Department of Biochemistry and Biophysics, Stockholm University, Solna, 171 65, Sweden; Division of Internal Medicine, Department of Clinical Sciences, Karolinska Institutet, Danderyd Hospital, 182 57 Danderyd, Sweden; Karolinska University Laboratory, Karolinska University Hospital, 141 86 Stockholm, Sweden; Department of Clinical Neuroscience, Karolinska Institutet, 171 77 Stockholm, Sweden

## Abstract

The COVID-19 pandemic has posed a tremendous challenge for the global community. We established a translational approach combining home blood sampling by finger-pricking with multiplexed serology to assess the exposure to the SARS-CoV-2 virus in a general population. The developed procedure determines the immune response in multiplexed assays against several spike (S, here denoted SPK), receptor binding domain (RBD) and nucleocapsid (NCP) proteins in eluates from dried capillary blood. The seroprevalence was then determined in two study sets by mailing 1000 blood sampling kits to random households in urban Stockholm during early and late April 2020, respectively. After receiving 55% (1097/2000) of the cards back within three weeks, 80% (878/1097) were suitable for the analyses of IgG and IgM titers. The data revealed diverse pattern of immune response, thus seroprevalence was dependent on the antigen, immunoglobulin class, stringency to include different antigens, as well as the required analytical performance. Applying unsupervised dimensionality reduction to the combined IgG and IgM data, 4.4% (19/435; 95% CI: 2.4%-6.3%) and 6.3% (28/443; 95% CI: 4.1%-8.6%) of the samples clustered with convalescent controls. Using overlapping scores from at least two SPK antigens, prevalence rates reached 10.1% (44/435; 95% CI: 7.3%-12.9%) in study set 1 and 10.8% (48/443; 95% CI: 7.9%-13.7%). Measuring the immune response against several SARS-CoV-2 proteins in a multiplexed workflow can provide valuable insights about the serological diversity and improve the certainty of the classification. Combining such assays with home-sampling of blood presents a viable strategy for individual-level diagnostics and towards an unbiased assessment of the seroprevalence in a population and may serve to improve our understanding about the diversity of COVID-19 etiology.

**One Sentence Summary:** A multiplexed serology assay was developed to determine antibodies against SARS-CoV-2 proteins in home-sampled dried blood spots collected by finger pricking.

## Introduction

SARS-CoV-2 emerged into the human population in Wuhan, China in late 2019, and the associated disease was termed COVID-19. At present, it has spread to almost every country in the world, infecting more than 10 million individuals, leading to at least 500,000 deaths. Accordingly, the WHO declared the pandemic to be a public health emergency of international concern. The virus is the seventh known human coronavirus. Four are known to circulate in the human population, typically causing common colds that occasionally advance to pneumonia. Prevalence of serum antibodies to these viruses can reach 90% or more in the population *(1)*.

Since many cases of SARS-CoV-2 infection are asymptomatic or present with mild disease, but still likely capable of spreading the infection, diagnostics has become vital in the understanding of the pandemic *(2)*. RT-PCR to detect viral RNA is important in a diagnostic context, is scalable to allow large scale testing but only detects ongoing infection. In contrast, serological assays that detect antibodies to viral antigens can reveal previous infection and thus are vital to understand the prevalence of the infection and for estimating population immunity. During coronavirus infection, patients typically develop strong antibody responses to the nucleocapsid protein and to the spike glycoprotein *(3)*. Indeed, the spike protein (here denoted SPK) has been one of the promising new targets for recently developed ELISA assays *(4, 5)*. The protein is a trimeric glycoprotein that can be divided by a protease cleavage site into membrane anchored S2 domain, and an N-terminal S1 domain, which includes the receptor binding domain (RBD). In addition, the nucleocapsid protein (NCP) is also being used in traditional ELISA assays. Multiplexed technologies that determine circulating antibodies against multiple analytes at the same time *(6)* have just recently begun to emerge for SARS-CoV-2 antigens *(7–9)*.

Serological testing has given estimates for seroprevalence rates in several populations, but often these are based only on specific inclusion criteria or sub-populations *(10, 11)*. Testing in blood donors has been suggested as a method to estimate the seroprevalence in presumably asymptomatic persons *(12)*. Medical laboratory investigations are based on sampling performed by health professionals while in point-of-care testing the sampling is done by the patients themselves. Point-of-care testing offers convenience by enabling self-sampling, but many of these tests do not fulfill the precision requirements *(13)*, as compared to ELISA assays *(14)*.

One way to overcome this could be to combine home-sampling and laboratory analysis. This becomes attractive in patients with chronic disease in need of continuous monitoring but also in situations, such as the ongoing COVID-19 pandemic, where measures are taken to limit the risk of infecting others. Dried blood spots based on collecting capillary blood is a well-established method screening neonates for in-borne disease or measuring antibodies against virus proteins *(15)*. However, when there is a need for precise measurements, the established technology based on punching a subsample out of a filter paper is not accurate enough *(16)*. In response to this need several devices providing a precise volume of dried blood have been developed *(17, 18)*.

This study addressed this challenge by combining volumetric home-sampling of blood with multiplexed analysis of antibodies against several different SARS-CoV-2 antigens. Following the development of the assay and workflow, the feasibility of comparing different antigens, sample types and serological methods was investigated. In addition, we set out to determine the seroprevalence in the Stockholm population during April 2020 by mailing out 2000 blood collection cards followed by the multiplexed analyses of the dried blood samples.

## Results

A translational workflow that combines home-sampling of blood with multiplexed serology analysis for antibody response to SARS-CoV-2 infections was applied as shown in **Fig. 1**. and with further details and explanations in **Fig. S1**.

**Fig 1.**
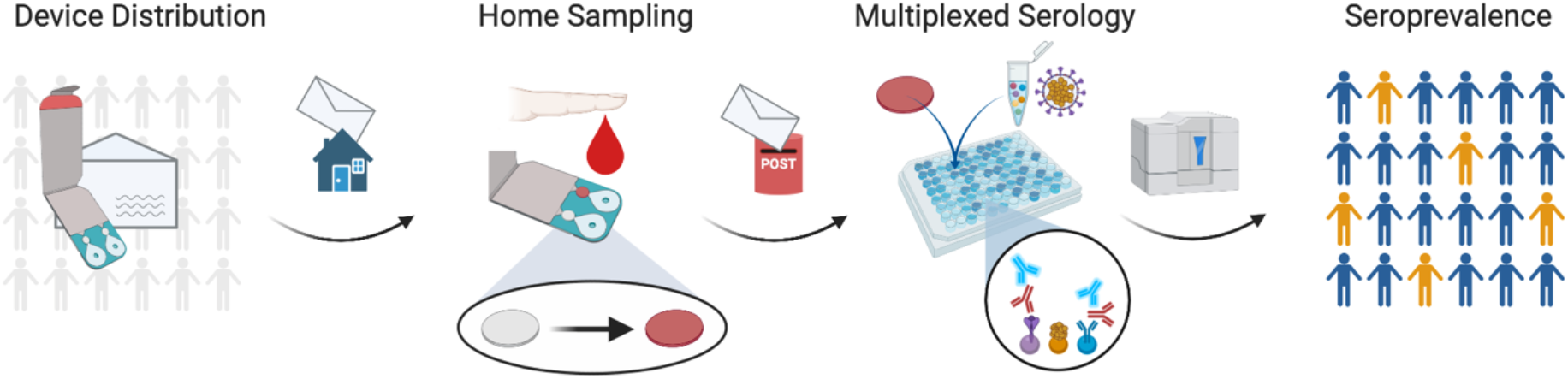
Translational approach for multiplexed serology in home-sampled dried blood spots. Blood collection devices were distributed by postal mail to collect blood samples from finger pricking at home. Cards were then mailed to the laboratory for multiplexed serological analysis. Data from antibody titers against multiple virus proteins was used to stratify individuals for seroprevalence.

First, a procedure to streamline the elution of proteins from dried blood samples (DBS) collected by finger pricking onto a volumetric microsampling device was developed. The DBS results were then compared with a commercial ELISA assays that detect relative IgG titers for the S1 and N proteins of the virus in venous plasma. Using the data set from this study, our in-house developed multiplexed assays utilizing suspension bead arrays (SBA) was benchmarked with data from the two single-analyte ELISAs. In parallel a total of 2000 blood collection devices were sent out to randomly selected inhabitants (age 20-74) in the urban area of Stockholm, Sweden. Sets of 1000 devices were sent during the first and last week of April 2020. Samples received prior to the end of April (study set 1) and the end of May (study set 2) were analyzed using the multiplexed SBA assays design to include multiple different virus antigens. Focusing on the stabilized prefusion spike protein (denoted SPK), a soluble fragment composed of the receptor-binding domain (RBD), and nucleocapsid protein (denoted NCP), we then assessed the seroprevalence via relative antibody titers.

### Assay development

An analytical pipeline was built on the Capitainer qDBS device that collects exactly 10 µl of blood after applying an unspecific volume of capillary blood obtained by finger pricking. Each device hosts two collection sites where a microchannel leads the blood onto a 6 mm paper disc where blood is then dried. The DBS samples were heat-inactivated prior to ejecting each disc into 96-well microtiter plates. The multiplexed serological procedure was built based on previous experience and protocols from profiling antibodies in serum or plasma *(19)(20)*. As described in further detail in the **Supplementary Text**, the method was evaluated for carry-over (**Fig S2**.), detectability (**Fig S3**.), and precision (**Table S1**.). On average, the intra-day CV was 13% (10%-15%) and the inter-day CV 18% (12%-22%), both being depended on the investigated antigen.

To perform multiplexed assays, different recombinant SARS-CoV-2 proteins and batches thereof were conjugated onto magnetic and color-coded beads (**Table 1**.). The acronyms SPK for the recombinant trimeric spike ectodomain, RBD for its receptor binding domain, and NCP for the nucleocapsid proteins were used. Two coupling chemistries were used to accommodate proteins stored in different buffers. With this set-up and liquid handling options for sample transfer and bead washing, 300 samples could be processed in about 300 min, and the sample incubation schemes were used for all studies and the detection of the immune response via either IgG or IgM. In summary, a workflow for the preparation of dried blood samples for serological analyses was developed and adapted to enable the determination of multiple analytes in each sample.

**Table 1A.** SARS-CoV-2 Proteins on beads.

| Protein | Acronym | Provider | Product ID/Note | Lot/Batch |
| --- | --- | --- | --- | --- |
| **Spike ectodomain** | SPK_01 | McInerney lab | Foldon, His-tag,<br>Strep-tag II | Batch 1 |
|  | SPK_02 |  |  | Batch 2 |
| **Spike ectodomain** | SPK_03 | Andréll lab | StrepIIHis-tag | Batch 1 |
| **Spike ectodomain** | SPK_04 | Andréll lab | His-tag | Batch 1 |
| **RBD** | RBD_01 | McInerney lab | FLAG-tag | Batch 1 |
|  | RBD_02 |  |  | Batch 2 |
| **RBD** | RBD_03 | McInerney lab | His-tag | Batch 1 |
| **RBD** | RBD_04 | Andréll lab | His-tag | Batch 1 |
| **Nucleocapsid** | NCP_01 | SinoBiological | 40588-V08B | LC14MC0309 |
| **Nucleocapsid** | NCP_02 | Elsässer lab | 2xStrep-tag | Batch 1 |
| **Nucleocapsid** | NCP_03 | Acro biosystems | NUN-C5227 | S34-2048F1-RB |
| **S1** | SPS_01 | SinoBiological | 40591-V08H | LC14MC1012 |

**Table 1B.** Other proteins.

| Protein | Acronym | Provider | Product ID/Note | Lot/Batch |
| --- | --- | --- | --- | --- |
| **SARS-CoV-1 S1** | SRS_01 | SinoBiological | 40150-V08B1 | LC14AP1505 |
| **MERS-CoV S1** | MRS_01 | SinoBiological | 40069-V08B1 | LC12AU0205 |
| **hCoV-NL63 S1** | NLS_01 | SinoBiological | 50600-V08H | LC14AP2005 |
| **EBNA1** | EBN_01 | Abcam | ab138345 | GR3235466-1 |

### Feasibility study

To benchmark our multiplexed serology assay procedure for DBS analysis, the data was compared to commercially available SARS-CoV-2 ELISA assays for the S1 and N proteins. In addition, antibody titers obtained from measuring IgG in blood collected by finger-pricking, venous blood prepared as EDTA plasma, as well as whole blood were compared. The samples for this feasibility study were collected by trained personnel and obtained from a local health care center together with some basic demographic information (**Table S2**.). The health care center had a high frequency of sick leaves during spring of 2020, hence, a bias for a higher prevalence of antibodies against the virus was expected. This pilot study set can therefore not be considered representative of the general population, but rather demonstrate how seroprevalence can increase in a local hotspot.

First, the commercial ELISA assays were used to compare plasma and DBS samples. Antibody titers were determined for the S1 and N protein as shown in **Fig. 2A**. In EDTA plasma, a very high seropositivity of 50% (25/50) was detected for the S1 antigen, and concordantly, 24 were seropositive for the N protein. The seropositivity overlapped for 92% (23/25) with two donors deemed positive only for one of the two protein. Additionally, DBS eluates from a subset of 38 volunteers were prepared and applied at the same theoretical amount as EDTA plasma. As shown in **Fig. 2B-C**, a high degree of concordance was found between DBS and plasma levels of IgG against the S1 and N proteins (rho > 0.9). Consequently, most of the DBS eluates were classified as the corresponding plasma samples. However, there were minor inconsistencies in the classification when either DBS or plasma levels were close to the designated cut-off levels. In general, this confirmed the suitability of DBS eluates for serological analysis.

**Fig 2.**
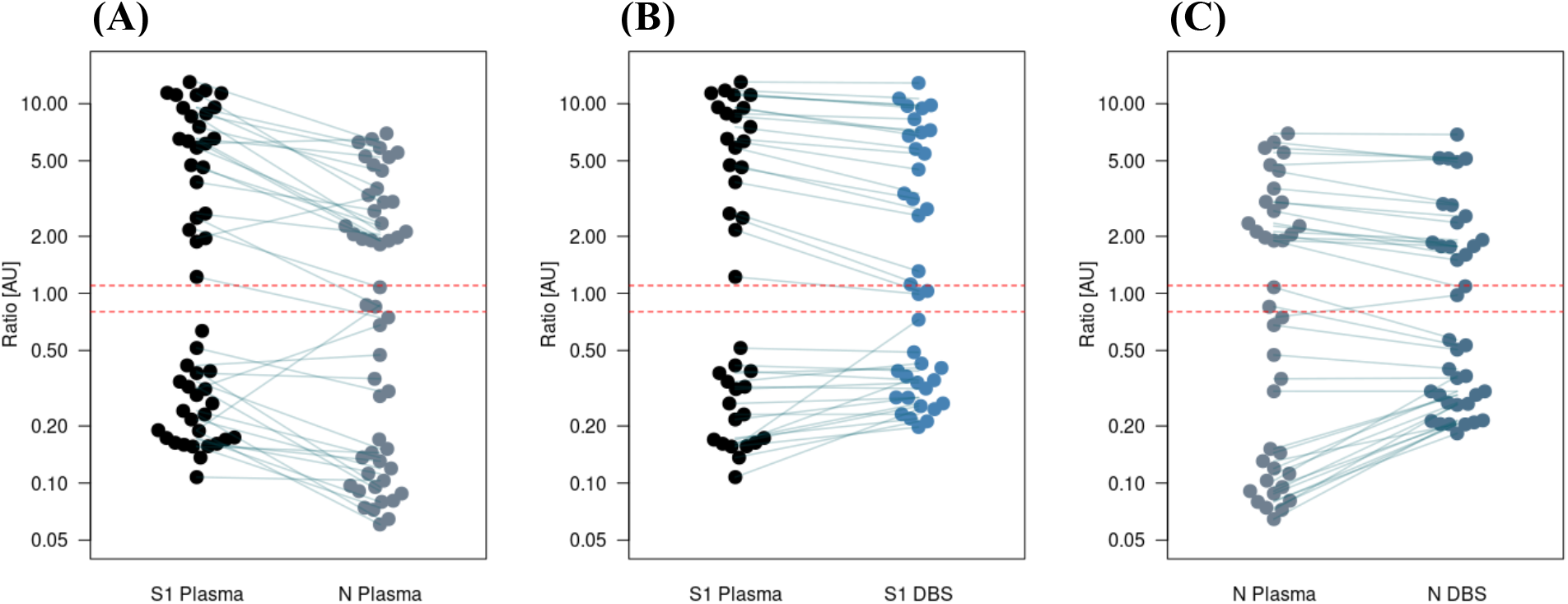
ELISA assays for S1 and N proteins in plasma and DBS eluates. **(A)** ELISA assays were performed on EDTA plasma from 50 subjects to detect titers against S1 (black) and the N protein (grey). Ratios between the internal controls are shown with the dotted lines indicating the recommended cut-off levels for seropositivity. Samples between the two red lines are borderline. **(B)** A comparsion between EDTA plasma (black) and DBS eluates (blue) for S1 ELISA with 38 subjects. **(C)** The same comparison is shown for the N protein in plasma (grey) and DBS eluates (blue), both showing a good concorance between the sample types.

Next, the DBS eluates and plasma samples were profiled using the multiplexed SBA assay workflow and results compared to the ELISA data. Since the SBA also contained three SARS-CoV-2 proteins from different sources, these were also compared to each other. An initial global analysis of the multiplexed data showed no distinct separation between EDTA plasma and eluates prepared from DBS (**Fig 3**). The latter included two types of DBS eluates, one prepared from finger pricked blood and the other prepared by applying whole blood from venous draws onto the DBS cards. Using S1 ELISA seropositivity, there was a clear separation of the two clusters in the PCA and UMAP analyses. There was only a single subject for which all three sample types did not group among the ELISA seropositive group. When investigating a possible reason, it turned out that the ELISA ratios of S1 = 1.19 and N = 0.75 were indicative for a classification as borderline. Indeed, the subject remained the only one that was deemed seropositive by ELISA for S1 but negative for N protein.

**Fig 3.**
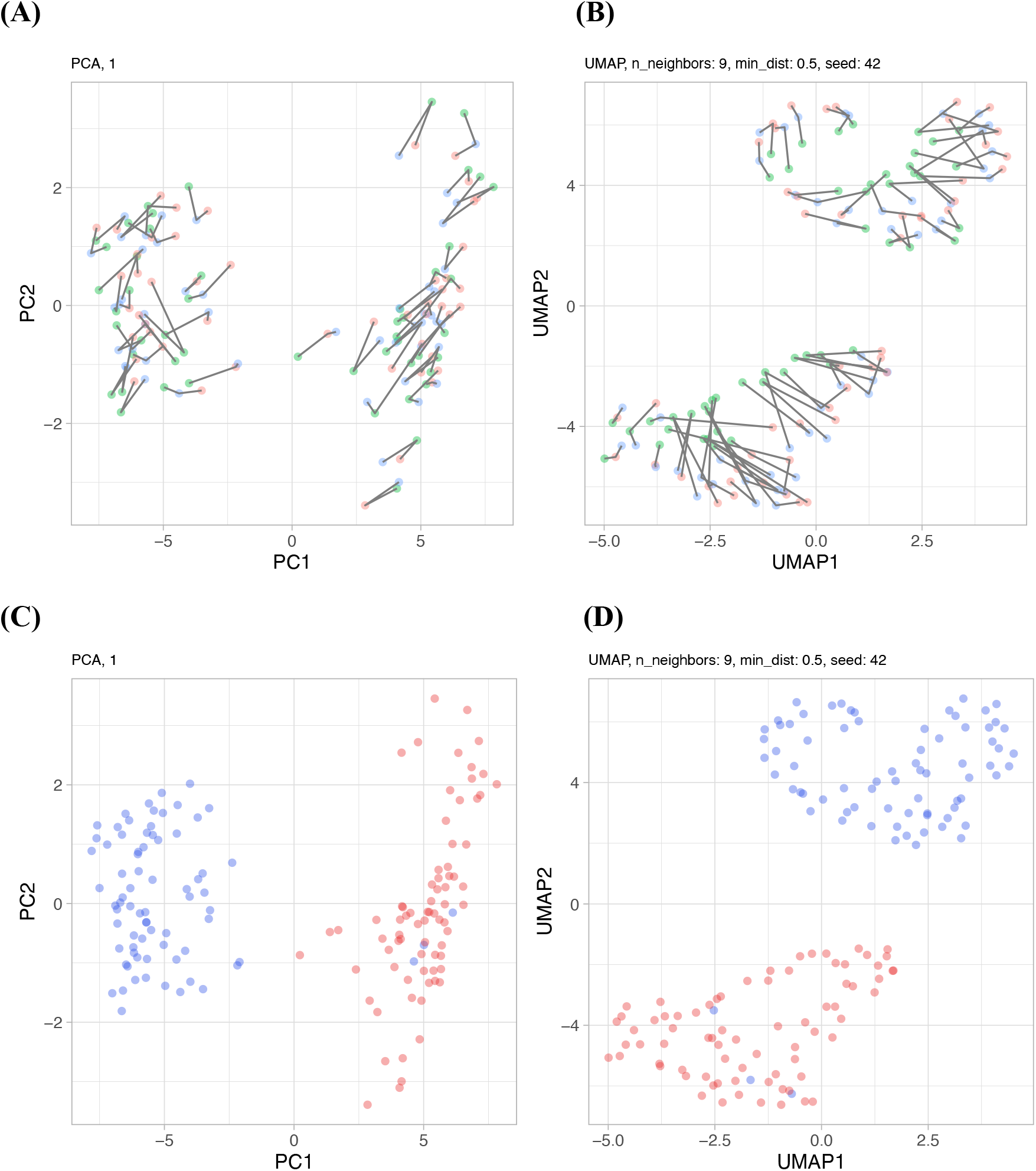
Multiplexed serology assays with plasma and DBS eluates. Dimensionality reduction of the multiplexed data was performed using PCA **(A, C)** and UMAP **(B, D)** for the viral antigens and internal controls. **(A-B)** PCA and UMAP analysis were colored based on plasma (green), capillary blood collected on DBS (cDBS, red), or whole blood from venous draws applied to the DBS cards (vDBS, blue). Samples from the same donors were connected by grey lines. **(C-D)** PCA and UMAP analysis were colored based on S1 ELISA seropositive (blue) and seronegative (red) subjects. There was a clear separation between seropositive and seronegative groups and samples from the same donor clustered in close distance.

As shown in **Fig. S4**. using both sample preparation types, there was supportive agreement in the classification between seropositive groups assigned by the S1 ELISA and the multiplexed serology data. No differences between the seropositivity groups detected for IgG levels or reactivity against the included control Epstein–Barr virus nuclear antigen 1 (EBNA1).

Correlation analyses were used to assess the degree of similarity between the ELISA and SBA data within plasma and DBS samples (**Fig. S5**.). For the NCP protein used in the ELISA and the three NCPs used in the SBA assays, there was a supportive similarity for DBS eluates (rho= 0.80±0.05) and plasma (rho= 0.76±0.06). The degree of concordance between S1 ELISA and SBA data for three SPK proteins was even higher in DBS eluates (rho= 0.86±0.005) as well as plasma (rho= 0.89±0.005). The different antigen preparations included in the SBA assay were also compared to obtain insights about the intra-assay consistency of reactivity levels. A high degree of concordance was determined between the three SPK proteins in DBS eluates (rho = 0.98±0.006) and plasma (rho = 0.99±0), between the four RBD proteins in (DBS: rho =0.91±0.06; plasma: rho=0.90±0.06), as well as between three NCP proteins (DBS: rho=0.86±0.06; plasma: rho =0.85±0.07). Taken together, these data indicated that the developed procedure performed on par with ELISA assays to identify seropositive and negative samples from plasma as well as DBS eluates, but the SBA’s multiplexing capacity provided further details the diversity of serological profiles for different antigens.

### Affinity proteomic analysis of DBS eluates

To assess possible differences in the overall protein composition of DBS and plasma samples as well as states of seropositivity, protein profiles obtained from multiplexed immunoassays for 92 circulating proteins were investigated. The levels of these proteins were ranked and compared in EDTA plasma for seropositivity and between in these paired EDTA plasma and DBS samples as summarized in the sheets of the **Supplementary Data File**. In these samples and with the used panel, 91 out of 92 proteins were detected in > 90% of the samples. When comparing the protein levels between samples grouped by their ELISA S1 seropositivity scores, there were no significant differences in (FDR < 0.01). The two top ranked proteins were the von Willebrand factor (vWF; p = 2.1E-03), which was more abundant in the seronegative group, and a cytokine from the tumor necrosis factor ligand family (TNFSF13B; p = 2.4E-03), with higher levels in the seropositive group. There were 33 proteins (36% of all tested) with significantly different abundance levels (FDR < 0.01) when DBS eluates were compared to matched EDTA plasma samples from 12 donors. Among these 33 hits, 26 proteins were more abundant in DBS eluates than in plasma. The proteins bleomycin hydrolase (BLMH; FDR = 1.83E-27), cystatin B (CSTB; FDR = 9.04E-25), proteinase 3 (PRTN3; FDR = 9.04E-25) and galectin 3 (LGALS3; FDR = 3.77E-24) were the three most differently abundant and elevated in DBS. For plasma, levels of interleukin-18-binding protein (IL18BP; FDR = 4.99E-06), C-X-C motif chemokine ligand 16 (CXCL16; FDR = 1.29E-06), collagen type I alpha 1 chain (COL1A1; FDR = 1.99E-05) and tissue factor pathway inhibitor (TFPI; FDR = 8.71E-03) were elevated compared to DBS eluates. There were only two proteins with significantly elevated levels in DBS collected by finger-pricking (cDBS) when compared with applying whole blood to the cards (vDBS). These were collagen type I alpha 1 chain (COL1A1; FDR = 3.11E-03) and integrin subunit beta 2 (ITGB2; FDR = 3.06E-02). The proteomics analyses of circulating proteins in DBS eluates support the integrity of these samples and additional assays can be performed to increase our understanding of the COVID-19 etiology.

### Longitudinal serology analysis

To investigate how IgG and IgM responses against the several different SARS-CoV-2 antigens changed over time and to assess changes in profiles from repeated sampling, a single PCR-positive donor collected blood on the DBS cards two weeks after a self-reported onset of COVID-19 symptoms. Blood was collected at five occasions over two weeks in intervals of about 3-8 days and until 30 days after symptom onset. Samples were analyzed within four weeks post collection with the multiplexed assay. As shown in **Fig. 4**. for IgG and IgM titers of several SARS-CoV-2 antigens, the shape of the longitudinal profiles depended on which immunoglobulin was detected.

**Fig 4.**
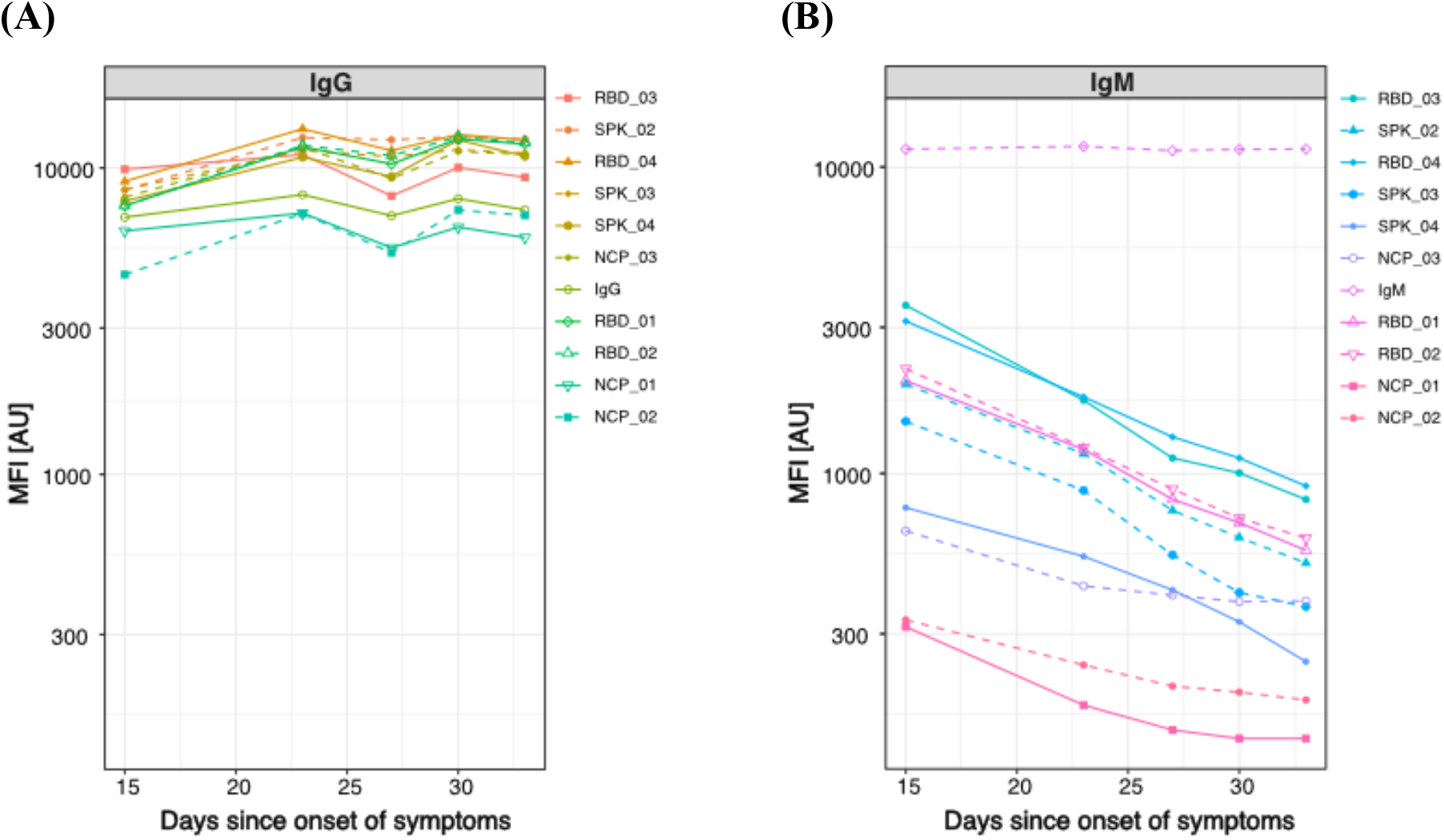
Longitudinal serology profiles. One PCR-positive donor collected blood on DBS cards at five occasions since self-reported onset of symptoms. Eluates from the DBS were analyzed for multiple virus proteins for titers of **(A)** IgG and **(B)** IgM. While IgG reactivity levels remained elevated throughout the sampling period, the levels of IgM against SARS-CoV-2 antigens declined over time. The data is presented as MFI values with each antigen presented in a unique color and symbol.

Linear regression was performed to extract and rank the changes in antibody titers. We found that levels of IgG (slope = 0.01) and IgM (slope = 0.003) remained unchanged during the sampling period. Combing the slopes from several RBD, SPK and NCP proteins, the IgG titers slightly increased with time (slope = 0.11±0.03) while a reduction of IgM titers was observed (slope = – 0.39±0.12). Titers for SPK (slope = –0.47) and RBD (slope = –0.45) decreased more than for NCP (slope = –0.22). The observed decline in IgM and stable IgG titers are in line with studies conducted in serum or plasma samples and further strengthen the suitability of the DBS-based multiplexed serology approach. The stable profiles obtained in this short-term longitudinal analysis also indicated the reproducibility of the workflow.

### Population study

A common challenge in assessing seroprevalence is the enrollment of representative participants and professional skills needed to collect blood samples. To minimize the possible bias of enrollment and to simplify the sampling procedure, the qDBS device and mail distribution of home-sampling kits were used to collect blood from the urban population in Stockholm, Sweden.

Two sets of 1000 home-sampling kits were sent out early and late April of 2020 to randomly selected individuals aged 20 to 74. As shown in **Table 2**., 55% of the sampling cards were received back and some had to be excluded due to incomplete disc loading or late return. Of the returned sampling cards, 82% of the participants succeeded to home-sample blood and provided at least one disc correctly filled with blood. The sampling dates were inferred from the signed consent forms (**Fig. S6A**.), and distribution of age-ranges as well as sex generally matched with those obtained from the population registers (**Fig. S6B**.). Differences in both compliance and sampling success rates were observed between sexes. In both sets about 13% self-reported fever under flu-like symptoms, and 22% listed symptoms related to issues with breathing, while > 50% of the participants reported no symptoms.

**Table 2.** Demographics of two population-based study sets.

|  | Study Set 1 | Study Set 2 | Overall |
| --- | --- | --- | --- |
| **Participation** |  |  |  |
| **Cards distributed** | 1000 | 1000 | 2000 |
| **Cards returned** | 529 (48.2%) | 568 (51.8%) | 1097 (54.8%)* |
| **Cards approved** | 435 (49.5%) | 443 (50.5%) | 878 (43.9%)* |
| **Sex** |  |  |  |
| **Female** | 239 (54.9%) | 244 (55.1%) | 483 (55.0%) |
| **Male** | 170 (39.1%) | 177 (40.0%) | 347 (39.5%) |
| **Other** | 1 (0.2%) | 0 (0%) | 1 (0.1%) |
| **Missing** | 25 (5.7%) | 22 (5.0%) | 47 (5.4%) |
| **Age Groups** |  |  |  |
| **20-29** | 67 (15.4%) | 68 (15.3%) | 135 (15.4%) |
| **30-39** | 77 (17.7%) | 97 (21.9%) | 174 (19.8%) |
| **40-49** | 74 (17.0%) | 91 (20.5%) | 165 (18.8%) |
| **50-59** | 87 (20.0%) | 64 (14.4%) | 151 (17.2%) |
| **60-69** | 66 (15.2%) | 64 (14.4%) | 130 (14.8%) |
| **70-74** | 39 (9.0%) | 40 (9.0%) | 79 (9.0%) |
| **Missing** | 25 (5.7%) | 19 (4.3%) | 44 (5.0%) |
| **Flu-like symptoms** |  |  |  |
| **No** | 221 (50.8%) | 246 (55.5%) | 467 (53.2%) |
| **Yes, mild** | 122 (28.0%) | 113 (25.5%) | 235 (26.8%) |
| **Yes, fever** | 58 (13.3%) | 58 (13.1%) | 116 (13.2%) |
| **Yes, severe** | 9 (2.1%) | 5 (1.1%) | 14 (1.6%) |
| **Missing** | 25 (5.7%) | 21 (4.7%) | 46 (5.2%) |
| **Breath symptoms** |  |  |  |
| **No** | 325 (74.7%) | 315 (71.1%) | 640 (72.9%) |
| **Yes** | 85 (19.5%) | 108 (24.4%) | 193 (22.0%) |
| **Missing** | 25 (5.7%) | 20 (4.5%) | 45 (5.1%) |

\* Related to total number of cards distributed

A primary focus was to determine the seroprevalence with IgG. For study set 1, titers were determined on two SPK, two RBD preparations, and one NCP protein. For study set 2, titers against three SPK, four RBD, and three NCP protein preparations were obtained. The distributions of the data for these antigens, other included proteins and controls can be found in **Fig. S7A-B**.

For each study set, different approaches were explored to calculate the seroprevalence levels. At first, the proportion of seropositive was determined by using a population-based model. The approach assumed that a majority of the analyzed samples did not contain any antibodies against the SARS-CoV-2 antigens. For each antigen, the peak of population titers served as the center point for defining a cut-off level and standard deviation (SD). For each antigen and study set, the resulting prevalence rates were then calculated and as shown in **Table 3A**. The results of the seroprevalence at this 6x SD cut-off ranged from 3% to 15% depending on the antigen and study set. Results for 3x SD are found in **Table S3**., and in the **Supplementary Data File** for all other antigens. The stated specificity and sensitivity levels were here based on the included and available PCR positive controls as well as negative controls collected prior to the outbreak. In summary, seropositive samples were identified in the population analysis using the dried blood samples.

**Table 3A.** Antigen-centric determination of IgG seropositivity. **Table 3A. Prevalence levels were calculated on 6x SD in each study set**. Numbers of samples above (Pos.) and below (Neg.) this cut-off are listed. Performance on sensitivity (Sens), specificity (Spec), as well as positive predictive value (PPV) were calculated using the included positive and negative controls. For set 1, there were 24 positive controls (4 unique plasma donors) and 45 negative controls (25 unique DBS blood donors). For set 2, there were 16 positive controls (2 unique DBS blood donors) and 50 negative controls (50 unique DBS blood donors).

| Antigen | Set | Prevalence | 95% CI | Pos. | Neg. | Sens | Spec | PPV |
| --- | --- | --- | --- | --- | --- | --- | --- | --- |
| NCP_01 | 1 | 7.36% | 4.90%- 9.81% | 32 | 403 | 100% | 100% | 100% |
| RBD_01 | 1 | 5.06% | 3.00%- 7.12% | 22 | 413 | 100% | 100% | 100% |
| RBD_02 | 1 | 6.21% | 3.94%- 8.47% | 27 | 408 | 100% | 100% | 100% |
| SPK_01 | 1 | 10.11% | 7.28%-12.95% | 44 | 391 | 100% | 96% | 71% |
| SPK_02 | 1 | 11.03% | 8.09%-13.98% | 48 | 387 | 100% | 98% | 83% |
| NCP_01 | 2 | 9.26% | 6.56%-11.95% | 41 | 402 | 100% | 98% | 85% |
| NCP_02 | 2 | 8.58% | 5.97%-11.19% | 38 | 405 | 100% | 96% | 74% |
| NCP_03 | 2 | 7.45% | 5.00%- 9.89% | 33 | 410 | 100% | 100% | 100% |
| RBD_01 | 2 | 5.87% | 3.68%- 8.06% | 26 | 417 | 100% | 100% | 100% |
| RBD_02 | 2 | 6.09% | 3.87%- 8.32% | 27 | 416 | 100% | 100% | 100% |
| RBD_03 | 2 | 10.16% | 7.34%-12.97% | 45 | 398 | 100% | 100% | 100% |
| RBD_04 | 2 | 10.61% | 7.74%-13.48% | 47 | 396 | 100% | 100% | 100% |
| SPK_02 | 2 | 8.13% | 5.58%-10.67% | 36 | 407 | 100% | 100% | 100% |
| SPK_03 | 2 | 11.06% | 8.14%-13.98% | 49 | 394 | 100% | 98% | 85% |
| SPK_04 | 2 | 11.51% | 8.54%-14.48% | 51 | 392 | 100% | 98% | 85% |

**Table 3B.** Combinatorial determination of IgG seropositivity.

| Seropositivity Criteria | Set | Prevalence | 95% CI | Pos. | Neg. | Sens | Spec | PPV |
| --- | --- | --- | --- | --- | --- | --- | --- | --- |
| **2xSPK** | 1 | 10.11% | 7.31%-12.92% | 44 | 391 | 100% | 98% | 83% |
|  | 2 | 10.84% | 7.94%-13.73% | 48 | 395 | 100% | 98% | 85% |
| **UMAP (PCR+ cluster)** | 1 | 4.37% | 2.46%- 6.27% | 19 | 426 | 100% | 100% | 100% |
|  | 2 | 6.32% | 4.05%- 8.59% | 28 | 415 | 100% | 100% | 100% |

SARS-CoV-2 is a lineage B betacoronavirus and groups together with SARS-CoV-1. To investigate antibody profiles against SARS-CoV-1 or other viruses such as MERS-CoV (lineage C betacoronavirus), human coronavirus NL-63 (alphacoronavirus), the S1 antigens representing MERS (MRS_01), SARS-CoV-1 (SRS_01) and NL-63 (NLS_01) were included in the second study set. At a 6x SD cut-off, none of these antigens could identify any of the included PCR-positive SARS-CoV-2 control samples (sensitivity = 0%). Profiles of MRS_01 and SRS_01 correlated with one another (rho = 0.55). None of the three antigens correlated with any of the SARS-CoV-2 antigens (rho < 0.3). Hence, these antigens were not informative regarding the determination of SARS-CoV-2 prevalence and provide a supportive indication for the specificity of SPK, RBD and NCP proteins.

Next, multiple antigens were combined to investigate seroprevalence levels. The UpSet visualization technique was used as it enables to aggregate combinations of different data, hence to find commonalities between seropositivity assignments obtained from different antigens. For each of the study sets, the combinations of different SARS-CoV-2 antigens are shown in **Fig. 5A-B**. when using IgG detection at a 6x SD cut-off level. Different combinations of the three antigens determined seroprevalence were observed and those involving any of the SPK proteins revealed the highest number of seropositive samples as compared to investigated RBD or NCP antigens. As shown in **Table 3B**., combining the scores from at least two of the included SPK protein (denoted 2xSPK) revealed prevalence levels of 10.1% in set 1 and 10.8% in set 2.

**Fig 5.**
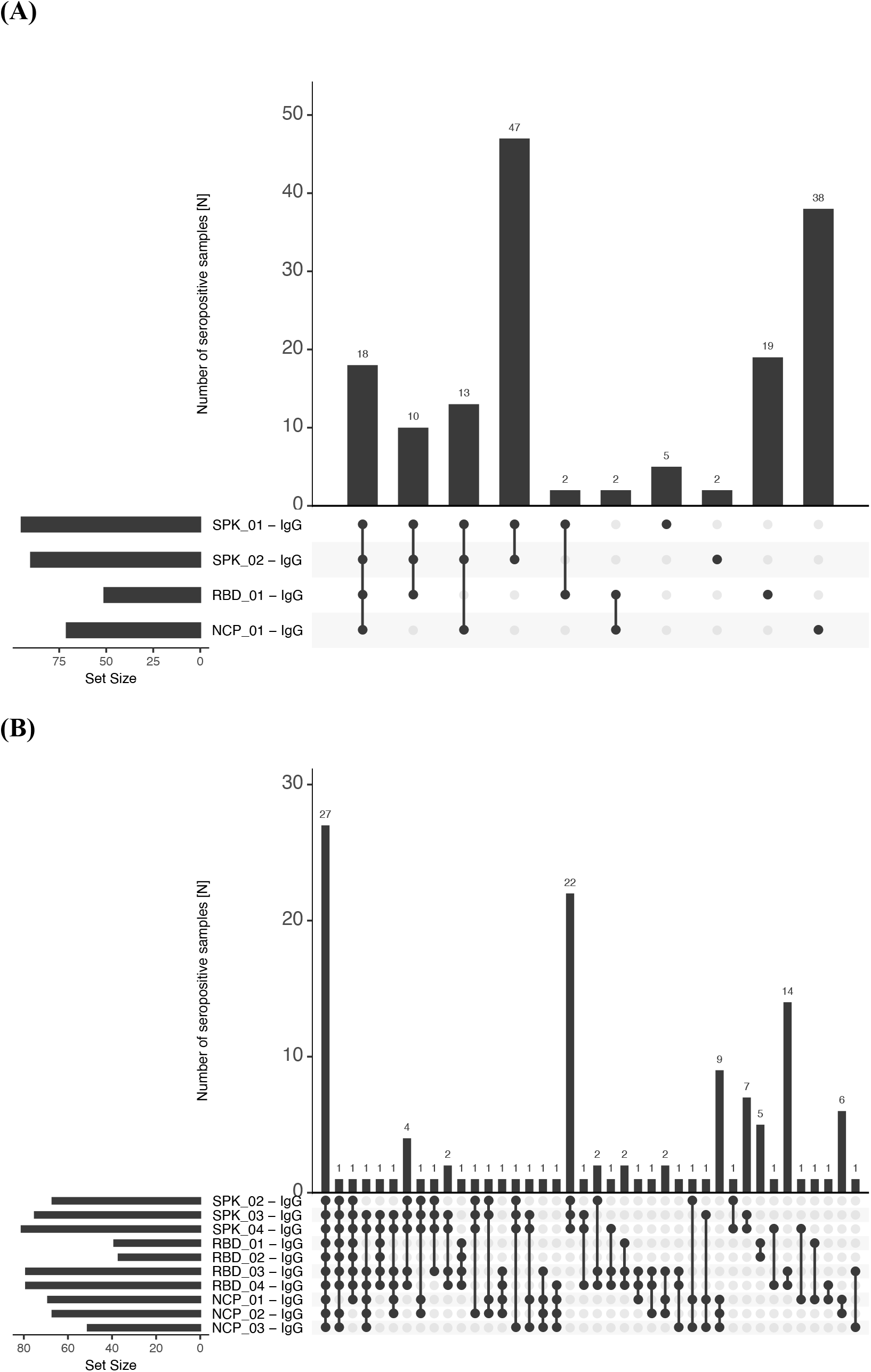
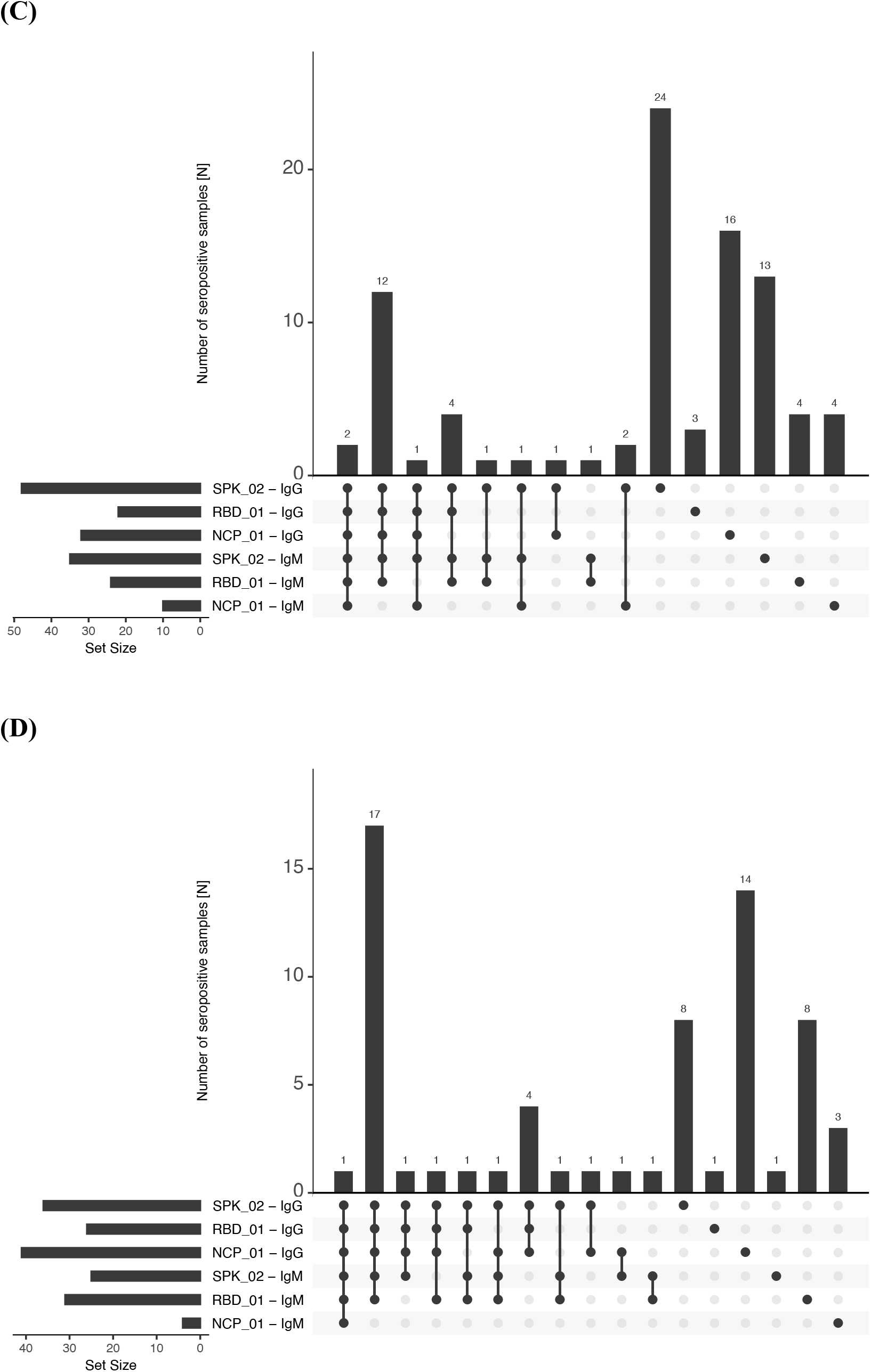
Combinatorial assessment of seroprevalence. Reactivity profiles obtained from different SARS-CoV-2 antigens were used to determine seropositivity in the study population. Intersections between the IgG-seropositivity scores are shown for the antigens used in **(A)** study set 1 and those included in **(B)** study set 2. The overlap between the IgG- and IgM-seropositivity scores are shown for three representative antigens in **(C)** for study set 1 and **(D)** for study set 2.

While most of the RBD-seropositive samples were also identified by SPK proteins, the scores assigned by NCP_01 were unique to 50% (16/32) of the samples in set 1 and 34% (14/41) in set 2. In **Fig. 5C-D**., the seropositivity scores obtained from both IgG and IgM were summarized to provide some possible insights into the time elapsed since infection. Of note, when compared with SPK and RBD proteins, there were very few samples with NCP-derived seropositivity when detecting IgM.

To obtain an alternative overview of seroprevalence in each study set, dimensionality reduction of the multiplexed serology data was performed using PCA and UMAP. As shown in **Fig. 6**. For both study sets, there were two sub-populations in the UMAP analyses when combining IgG and IgM data. There were 19/435 samples (4.4%) in study set 1 and 28/443 samples (6.3%) in set 2 that clustered with the included PCR-positive controls in this analysis. The majority of the remaining samples clustered with the included negative controls. Since the positive controls were taken from convalescent donors with expected strong immune response against the virus, the profiles from the 47 study participants (representing 5.4% of all samples) were expected to differ in their serology profiles for many of the virus proteins.

**Fig 6.**
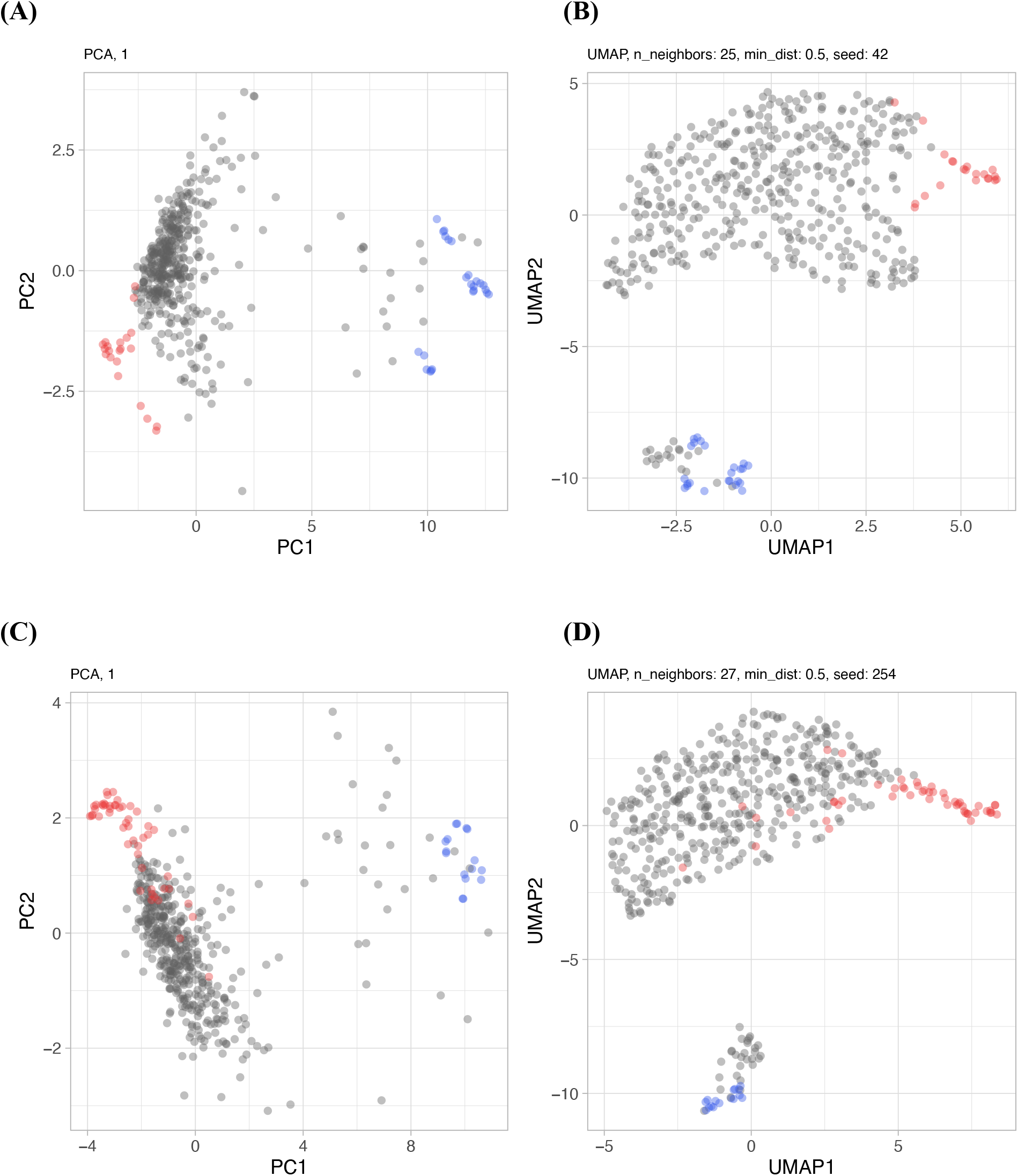
Global analysis of multiplexed serology data. Two dimensionality reduction techniques identified patterns of seropositivity in (**A-B**) study set 1 and (**C-D**) study set 2 by combining the IgG and IgM reactivity profiles. The color codes indicate the samples from the population (grey), negative controls collected prior to 2020 (red) and PCR-confirmed positive controls (blue). For PCA **(A**,**C)** and UMAP **(B, D)** viral antigens for study set 1 included SPK_01, SPK_02, RBD_01, RBD_03, NCP_01, and for study set 2 SPK_02, RBD_01, RBD_02, RBD_03, NCP_01.

Matching these samples with sex, age-range and flu-like symptom seronegative samples (see affinity proteomics analysis below) there were expected differences in profiles from the SPK, RBD and NCP proteins with IgG detection, while NCP did not differ between these groups when detecting with IgM **(Fig. S8**. for IgG and **Fig S9**. for IgM). There were no differences between these groups in terms of their IgG or IgM titers as well as for those found for EBNA-1.

Consequently, the relationship between the demographic information and groups of participants classified as seropositive or seronegative was investigated. The criteria involving at least two SKP proteins (2xSPK) as well as the clusters identified by UMAP analysis (see above) were chosen. As shown in **Table 4**., both classification approaches found significant differences (P < 0.05) in the proportional distributions.

**Table 4.** Association between seropositivity rates and demographic information. Nominal p-values for the comparison of seropositivity within the age and flu-like symptom groups were conducted by Wilcoxon’s rank sum test. For the comparisons between sex and breathing symptoms groups, a Fisher’s exact test was used. OR: Odds Ratio. N/A: Not available.

|  | 2xSPK |  |  | UMAP |  |  |
| --- | --- | --- | --- | --- | --- | --- |
|  | Positive | Negative | P-value (OR) | Positive | Negative | P-value |
| Samples |  |  |  |  |  |  |
| Groups | 92 (10.5%) | 786 (89.5%) |  | 47 (5.4%) | 831 (94.6%) |  |
| Sex |  |  |  |  |  |  |
| Female | 54 (58.7%) | 429 (54.6%) | 0.9<br>(1.05) | 28 (59.6%) | 455 (54.8%) | 0.8<br>(1.06) |
| Male | 37 (40.2%) | 310 (39.4%) |  | 19 (40.4%) | 328 (39.5%) |  |
| Other | 0 (0%) | 1 (0.1%) |  | 0 (0%) | 1 (0.1%) |  |
| Missing | 1 (1.1%) | 46 (5.9%) |  | 0 (0%) | 47 (5.7%) |  |
| Age group (years) |  |  |  |  |  |  |
| 20-29 | 24 (26.1%) | 111 (14.1%) | 0.1<br>(N/A) | 15 (31.9%) | 120 (14.4%) | 0.006<br>(N/A) |
| 30-39 | 14 (15.2%) | 160 (20.4%) |  | 10 (21.3%) | 164 (19.7%) |  |
| 40-49 | 16 (17.4%) | 149 (19.0%) |  | 8 (17.0%) | 157 (18.9%) |  |
| 50-59 | 17 (18.5%) | 134 (17.0%) |  | 6 (12.8%) | 145 (17.4%) |  |
| 60-69 | 16 (17.4%) | 114 (14.5%) |  | 7 (14.9%) | 123 (14.8%) |  |
| 70-74 | 4 (4.3%) | 75 (9.5%) |  | 1 (2.1%) | 78 (9.4%) |  |
| Missing | 1 (1.1%) | 43 (5.5%) |  | 0 (0%) | 44 (5.3%) |  |
| Flu-like symptoms |  |  |  |  |  |  |
| No | 33 (35.9%) | 434 (55.2%) | < 0.001<br>(0.40) | 8 (17.0%) | 459 (55.2%) | < 0.001<br>(0.15) |
| Yes, mild | 27 (29.3%) | 208 (26.5%) |  | 15 (31.9%) | 220 (26.5%) |  |
| Yes, fever | 27 (29.3%) | 89 (11.3%) |  | 20 (42.6%) | 96 (11.6%) |  |
| Yes, severe | 4 (4.3%) | 10 (1.3%) |  | 4 (8.5%) | 10 (1.2%) |  |
| Missing | 1 (1.1%) | 45 (5.7%) |  | 0 (0%) | 46 (5.5%) |  |
| Breathing symptoms |  |  |  |  |  |  |
| No | 60 (65.2%) | 580 (73.8%) | 0.01<br>(0.54) | 28 (59.6%) | 612 (73.6%) | 0.007<br>(0.42) |
| Yes | 31 (33.7%) | 162 (20.6%) |  | 19 (40.4%) | 174 (20.9%) |  |
| Missing | 1 (1.1%) | 44 (5.6%) |  | 0 (0%) | 45 (5.4%) |  |

In both seropositive groups with antibodies against SARS-CoV-2 proteins, there were more participants reporting fever or severe flu-like symptoms as well as symptoms related to cold-related issues with breathing. The 2xSPK approach found more asymptomatic and presumably mild cases, and both classification approaches found 4 subjects reporting severe symptoms. While no difference in the age distribution was found when using at least two SPK proteins, it was when using the UMAP classification that slightly more seropositive participants were younger than the seronegative group.

Finally, affinity proteomics analyses in DBS eluates were performed to search for any differently abundant proteins in relation to a previous SARS-CoV-2 infection. Three protein panels targeting a total of 276 proteins were chosen due to the lower protein concentration of the DBS eluate as compared to plasma. A set of 42 individuals deemed seropositive by the UMAP analysis were selected and compared against 42 age-range, sex and flu-like symptoms individuals that were seronegative. Only 16 out of 276 proteins (5.8%) could not be detected in > 10% of the samples, which indicates the suitability of the eluates for this analysis. While there were no significant differences (FDR < 0.01) between the groups in relation to the seroprevalence, there was a trend that many of the analyzed proteins were detected to be more abundant in the seropositive group (**Fig. S8**., **Supplementary Data File**). The three top hits were the acute phase liver protein mannose binding lectin 2 (MBL2; p = 2.1E-04; FDR = 0.06), glycoprotein Ib alpha chain (GP1BA; p = 2.3E-03) found in lymph nodes and blood granulocytes, as well as and the cell adhesion molecule L1-like protein (CHL1; p = 2.5E-03; FDR > 0.1), which is expressed in B-cells.

## Discussion

Our study used a translational serology approach to determine the prevalence of antibodies against several of the SARS-CoV-2 proteins in blood from finger prick samples. By utilizing a dedicated home-sampling system in combination with multiplexed immunoassay systems, we profiled dried blood samples from anonymous study participants in Stockholm, Sweden.

The collection of dried blood is a well-established procedure to obtain samples for a variety of clinical biomarker analyses. During the 2020 COVID-19 pandemic, sampling by laypeople outside the hospital and health care setting was considered to be particularly attractive as it reduced the burden of health care centers to sample patients not requiring hospital care. It also reduced the need for participants or patients to leave their homes and travel to these centers to give a blood sample. The latter is a noteworthy advantage considering the possible bias in studies where participation by invitation or selection *(21)*, while some individuals might not feel comfortable leaving their homes during a pandemic. This includes members of particular risk groups who should reduce the risk of exposures, but also those that were asymptomatic to COVID-19 or unwilling to participate. However, self-sampling poses some challenges, primarily concerning the lack of common experience in sampling blood via finger pricking and ensuring that the quality of the sample is suitable for laboratory testing. Even though the success rate of self-sampling was 82% for those sending back the cards, incomplete blood collection was occasionally observed, thus the components of the sampling kits as well as the instructions can be improved. In the presented study design, there was a risk for low compliance rates. Nonetheless, we received 55% of 2000 cards back and used 44% of the 2000 distributed cards for serological assays. Dried blood spots can offer biosafety benefits, particularly when volume-controlled sampling and simplified handling of these are made possible. These are due to small sample volumes being obtained, samples having dried for a few days between sampling and measurement, plus heat treatment and detergent elution being applicable to reduce the risk of infection. Further, samples can be shipped and stored at room temperature and thus do not require bulky dry ice packaging and freezer space.

Our study was designed to mail out collection kits to random individuals in Stockholm to reduce possible bias due to health, travel or socioeconomic status. The study was limited to Stockholm, which is the largest urban area in Sweden with approximately 2 million inhabitants, and the most COVID-19-affected area in the country with relatively high death rates. During the period of our study, there were approximately 4500 confirmed cases diagnosed early April 2020 (as per April 12) and the number increased to nearly 10,000 within four weeks (as per May 10) *(22)*. Participants were invited by regular mail to self-report limited information about their health condition during the previous months in relation to flu-like symptoms including fever, coughing and breathing difficulties. None of the participants in the population studies reported PCR positive status for SARS-CoV-2. We chose to send out sampling kits during the beginning and end of April to compare if and how seroprevalence would change during that time. As exemplified by the UMAP analysis, we found a minor increase in seroprevalence during this short period.

However, there are some limitations to the chosen design and approach. First, anonymized and population-based studies are more attractive but such a design limits the possibilities to conduct follow-up analyses. Other studies using multiplexed assays may therefore investigate the dynamics of antibody titers, such as IgM, and determine if particular antigens become dominant over time. Secondly, minimal and self-reported information limits the possibilities to draw further and more general conclusions about COVID-19, as both types of information can be biased by completeness and subjectivity of the reported scales. Our study can be expanded beyond Stockholm to more remote areas with greater distances to health care centers and include more than the analyzed 1000 DBS samples to increase the power and study the seroprevalence in different regions of Sweden and other countries.

The investigation studied the antibodies as well as proteins in DBS eluates from discs having dried blood from a volume of 10 µl. These 3D matrices fitted into regular 96-well microtiter plates, hence opened up the possibility to process samples in a high-throughput format. Pre-analytical sample handling was minimal. The protocol included the addition of elution buffer to the blood discs, incubation, and centrifugation to prepare the eluates from the cell-free supernatants for the analyses. All steps were compatible with well-established protocols for antibody profiling with serum or plasma samples. Since whole blood consists of about 50% cell free fluid, we used this approximation to calculate the dilution factors for our assays. While further optimization of the sample preparation is possible, the addition of Tween20 and protease inhibitor cocktails were sufficient to obtain 70 µl of eluate that contained 350 µg of protein. This eluate contained about 70% of the theoretical 500 µg of plasma proteins in 10 µl of blood from the discs. The main utility of the protocol was to determine circulating levels of IgG and IgM, which are all abundant and stable blood proteins. However, as we also demonstrate by the multiplexed analysis of other circulating proteins, the DBS samples are also well suited for other types of proteomics assay.

Multiplexed immunoassays were used in order to determine the concentration of antibodies against several different antigens at the same time. The SBA platform offers a versatile technology to combine different types of baits and analytes into one assay in order to compare these under the identical conditions. Further, different sources or batches of the same antigens can be compared, including the effects of different epitope tags and protein coupling chemistries. This allowed us to internally validate the serological profiles and increase the certainty of the determined prevalence, but also demonstrated robustness of the assay and reproducibility when antigens were obtained from different sources. Since the DBS eluates were a limited resource the current protocol only allowed for four separate assays to be performed per disc. While further reduction in sample volume can increase the number of assays beyond IgG and IgM, it was obvious that multiplexing these analyses became even more important to scale and maximize the amount of information collected per sample. Some of the challenges for multiplexed assays are to adjust the conditions so that several different parameters can be determined with a comparable performance. This can be one of the reasons why some antigens outperformed others. Nonetheless, this mostly relates to the dilution of the sample and the risk that some analytes are below the limit of detection while others have reached saturation levels. It is therefore likely that the assays for some of the antigens did not perform as well as in single-analyte assays, or when using other sample types, coupling conditions, assay protocols, and immunoassay platforms.

Besides the technical and design aspects discussed above, we found discrepancies between the immune activities against the most commonly used SARS-CoV-2 antigens denoted SPK, RBD and NCP. Each of the three antigens offers possibilities and challenges. The main neutralizing antibody response is mounted against the viral spike protein, and especially against its subdomain, the RBD. While reactivity against RBD was always accompanied by reactivity against SPK, not all SPK positive individuals showed RBD reactivity. Accordingly, the SPK protein was found to reveal the highest seropositivity rates, which was mostly complementarity for both IgG and IgM detection. Reactivity towards RBD was common for both IgG and IgM, and some donors were only seropositive with IgM. Interestingly, there were less IgM-seropositive samples when using the NCP proteins. These observations could indicate that the NCP proteins might possibly be informative for earlier phases of the infection, or represent cross-reactivities from previous infections with other coronaviruses. More data from longitudinal studies are needed to validate these hypotheses. Nonetheless, multiplexing allowed us to include several antigens from the virus as well as other related viruses. There are indeed other alphacoronaviruses (HCoV-229E and HCoV-NL63) and lineage A betacoronaviruses (HCoV-OC43 and HCoV-HKU1) as well as SARS-CoV-1 to be considered. The profiles from the included human coronavirus NL63, MERS-CoV and SARS-CoV-1 revealed a different distribution of reactivity profiles than the SARS-CoV-2 antigens (**Fig. S7**.). When assessing their performance to detect SARS-CoV-2 antibodies with the included positive and negative controls (**Supplementary Data File**), their sensitivity was indeed 0%. The observed reactivities were therefore more likely due to cross-reactivity from antibodies against the SARS-CoV-2 proteins.

While assessing the seroprevalence, either via the reactivity profiles of individual antigens or via combination of these, we found that one of the current main challenges for each platform is to use the appropriate numbers of negative and positive controls. With the assumption that seroprevalence was < 50% for both population-based study sets, we expected the reported reactivity levels from the seronegative samples to be within the lower reactivity range. Hence, we chose these sub-populations as being negative for SARS-CoV-2 infection. When increasing the stringency of the cut-off, the number of false positives naturally decreased while some true positive cases might be lost. Even though we reported sensitivity and specificity levels of 100%, we do acknowledge the low number of positive controls and that further validation with more samples, other confirmed viral infections, as well as a range of mild and severe COVID-19 patients are needed to assess the true performance of the approach. A current hurdle for utilizing our translational workflow is therefore to find or even establish appropriate biobanks that host a diverse set of samples from PCR or antibody positive persons with the different types of viruses.

## Materials and Methods

### Samples and Sampling

#### Feasibility study

Venous as well as capillary blood samples were collected from volunteers (N=50) among personnel at a healthcare center in Stockholm between May 14-18, 2020 by a trained phlebotomist. Venous blood samples (two per donor) were collected through venipuncture into EDTA blood collection tubes (K2E K2EDTA Vacuette tube, #454410, Lot# A19104MX, Greiner Bio-One) and capillary blood samples were obtained by finger-pricking using a contact-activated lancet (BD Microtainer #366594, BD) and applying blood droplets onto a quantitative DBS sampling card (qDBS, Capitainer AB, Stockholm, Sweden) according to the supplier’s instructions. After blood collection, one of the venous blood tubes was centrifuged and the blood plasma was collected into a separate tube. Both the plasma sample and the other blood tube was stored at −20 °C until further use. The qDBS cards were stored at room temperature until being heat treatment prior to extracting the blood-filled discs.

#### Population study

Capillary blood samples from the general population were obtained by cold-mailing home-sampling kits to 2000 randomly selected individuals (20-74 years old) in metropolitan Stockholm (**Table S4**.) during April 2020. Home-sampling kits were mailed in standard C4 envelopes containing the kit with a contact-activated lancet (BD Microtainer #366594, BD), quantitative DBS sampling card (qDBS, Capitainer AB, Stockholm, Sweden), return pouch (Capitainer AB, Sweden), alcohol swab, gauze and band-aid, as well as an information letter, questionnaire, consent form, C5 prepaid return envelope, and an instruction sheet for home-sampling (MM20-009-01, Capitainer AB, Sweden). Individuals who volunteered to participate in the study were asked to perform self-sampling according to the instructions and return the filled sampling card, questionnaire and consent form in the prepaid return envelope by regular mail. The samples were analyzed within 3-4 weeks after receiving the last blood cards.

#### Positive control samples

EDTA plasma samples from four COVID-19-convalescent and PCR-confirmed individuals were obtained from Karolinska university hospital in Huddinge. All participants had recovered from a PCR-verified COVID-19 for at least 2 weeks. Ten µl of plasma were loaded directly onto each disc of DBS cards. In addition, we obtained capillary blood samples from five COVID-19-convalescent and PCR-confirmed individuals who volunteered to donate blood after hospital discharge and using the home-sampling procedure as above. Further to this, one COVID-19-convalescent and PCR-confirmed individual volunteered to donate capillary blood samples every 3-8 days during a three weeks period. Samples from ELISA and PCR-positive participants from the feasibility study were used as additional controls.

#### Negative control samples

As negative controls for set 1, we used 25 DBS samples from venous blood donors. Samples were collected from anonymous donors prior to 2020 and purchased from Blodcentralen (Region Stockholm). For set 2, we used 44 capillary blood DBS samples collected in a biobank from patients before 2019. In addition, we used commercially available EDTA plasma from a pool of anonymous healthy males and females (#HMPLEDTA2, Lot# BRH1176237, Seralab) that had been purchased prior to the COVID-19 outbreak and was stored at –80 °C until use.

All cards were barcoded and stored at room temperature until use, or as stated otherwise. All collected DBS samples were discarded after the study was completed. All blood donors gave informed documented consent. The study was approved by the regional ethical board (EPN Stockholm, Dnr 2015/867-31/1) and the Swedish Ethical Review Authority (EPM, Dnr 2020-01500). Use of biobanked controls samples was approved by the Swedish Ethical Review

Authority (Dnr 2020-02483). At Karolinka University Hostipal in Huddinge, corona serology testing as part of a convalescent plasma donation study was approved by the National Ethical Review Agency of Sweden (Dnr 2020-01479) and all participants provided written informed consent.

### Protein production

Recombinant proteins were either obtained from commercial providers or produced by the independent labs as follows and as summarized in **Table 1**. The following acronym codes were used for the different proteins and batches: For the spike ectodomain we used SPK, for the receptor binding domain we used RBD, and for the nucleocapsid proteins we used NCP.

#### Spike ectodomains

The McInerney lab obtained the plasmid for the expression of the SARS-CoV-2 prefusion-stabilized spike ectodomain from Wrapp et al *(23)*, as a gift from Jason McLellan at University of Texas, USA. The plasmid encoding the SARS-CoV-2 spike ectodomain followed by T4 fibritin trimerization motif, a 8xHIS tag and a StrepII-tag was used to transiently transfect FreeStyle 293F cells using FreeStyle MAX reagent (Thermo Fisher). The S1 ectodomain was purified from filtered supernatant on Streptactin XT resin (IBA Lifesciences), followed by size-exclusion chromatography on a superdex 200 in 5 mM Tris pH 8, 200 mM NaCl, and rebuffered into PBS before coupling to beads. The protein was produced on different dates as two batches following the same protocol, hence denoted SPK_01 and SPK_02.

The Andréll lab produced two spike ectodomain constructs. The Sfoldon-His-StrepIIHis protein (SPK_03) is the same construct as SPK_01. A second spike trimeric ectodomain (SPK_04, Sfoldon-His) was generated using a plasmid provided as a kind gift from John Briggs and Andrew Carter at Laboratory of Molecular Biology MRC, UK that were a modified version of SPK_01/03.

Expi293 were transiently transfected with SPK_03 or SPK_04 using PEI transfection reagent (# 23966, Polysciences). After 72 h post-transfection, the supernatant was cleared and Spike ectodomain purified on Ni-NTA resin (#88221, Thermofisher). For SPK_04 protein the Ni-NTA step was followed directly by size-exclusion chromatography on a Superose 6 gel filtration column in 20 mM HEPES, 200 mM NaCl. For SPK_03 protein the Ni-NTA step was followed by purification on Strep-Tactin XT resin (#2-4010-010, IBA) prior to gel filtration on a Suprose 6 gel filtration column in 20 mM HEPES, 200 mM NaCl.

#### RBD proteins

The McInerney lab prepared the two RBD constructs termed RBD_01 and RBD_03. RBD domain RVQ-VNF of SARS-CoV-2 (RBD_01) was cloned upstream of an enterokinase cleavage site and a human FC. This plasmid was used to transiently transfect FreeStyle 293F cells using the FreeStyle MAX reagent and this FC fusion was purified from filtered supernatant on Protein G Sepharose (GE Healthcare). The protein was cleaved using bovine enterokinase (GenScript) leaving a FLAG-tag on the C-terminus of the RBD. Enzyme and FC-portion was removed on His-Pur Ni-NTA resin and Protein G sepharose respectively, and the RBD was purified by size-exclusion chromatography on a Superdex 200 in 5 mM Tris pH 8, 200 mM NaCl. This protein was termed RBD_01 and a second batch, produced in the same way, but on a different day, was termed RBD_02. For the second variant, the RBD domain RVQ-QFG (RBD_03) was cloned upstream of a Sortase A recognition site and a 6x His-tag, and expressed in FreeStyle 293F cells as above. RBD_03 was purified from filtered supernatant on His-Pur Ni-NTA resin, followed by size-exclusion chromatography on a Superdex 200.

The Andréll lab prepared RBD_04 by using the RBD-His plasmid obtained from BEI resources NR52309 *(5)*. Expi293 cells were transiently transfected with RBD_04 using using PEI transfection reagent (# 23966, Polysciences). After 72 h post-transfection, the supernatant was cleared and RBD_04 purified on Ni-NTA resin (#88221, Thermofisher) followed by size-exclusion chromatography on a Superdex 200 gel filtration column in PBS.

#### NCP protein

The Elsässer lab prepared the nucleocapsid protein denoted NCP_02. The mammalian expression plasmid pLVX-EF1alpha-nCoV2019-N-IRES-Puro used for mammalian expression plasmid was a kind gift from Nevan Krogan lab at UCSF. HEK293T cells were transfected using PEI. Cells were harvested 60 h post-transfection and NCP protein affinity purified similar as described elsewhere *(24)*: cells were washed with PBS and lysed in 50 mM Tris, 150 mM NaCl, pH 7.4, 1 mM EDTA, 0.5% NP-40 supplemented with 1x complete protease inhibitor (#11873580001, Roche). Lysate was cleared by centrifugation and incubated with 20 µl 5% Strep-Tactin bead suspension (2-4090-002, IBA), 30 min on ice. The resin was washed 3x with 100 mM Tris/HCl, pH 8.0; 150 mM NaCl; 1 mM EDTA, 0.05% NP40 and eluted in the same buffer supplemented with 50 mM biotin.

### Experimental study design

#### Feasibility study

For the ELISA analysis, EDTA plasma (N=50) and DBS eluates prepared from finger pricked sampling (N=38) were analyzed together as described below. The multiplexed serology performed with the SBA assays were conducted in duplicate. The latter assay analyzed EDTA plasma (N=50), DBS eluates of whole blood collected by finger pricking (cDBS, N=50) as well as from DBS eluates of whole blood prepared by applying blood to the cards that had been collected from venous draw (vDBS, N=50).

#### Population study

Assuming that DBS cards were delivered by mail without any correlation with age or sex, one disc from each card was transferred into 96-well plates without additional randomization. Each 96-well plate was filled with 80 discs.

For the first set of population samples (Study Set 1, N=435), each 96-well assay plate had two empty filter-discs, two wells with assay buffer only, four discs from negative controls prepared from the pool of EDTA plasma collected prior 2019, four negative controls consisting of dried blood spots collected before the outbreak, and four positive controls in form of EDTA plasma samples applied to discs from COVID-19-convalencent donors. One well per plate was left empty for plate identification and orientation. The four control wells with healthy plasma had the same (pooled) sample, which provided data about reproducibility.

For the second set of population samples (Study Set 2, N=443), each 96-well assay plate had two wells with assay buffer only and two discs from two PCR-positive individuals that were present in all assay plates. In addition, the second discs of eight previously analyzed subjects from study set 1 and second discs of four individuals from the feasibility study were added. We also included and reanalyzed ten DBS eluates prepared for Study Set 1 as well as ten DBS eluates prepared for this study set.

### Sample preparation

First, the blood sampling cards were heated at 56°C for 60 min in an oven (UN55m, Memmert GmbH) in sets of 50. Each card was visually inspected to determine if at least one paper disc was correctly filled with blood. From each card deemed successful, one paper disc was ejected using a semi-automated card-punching apparatus (qDBS Card Puncher, Capitainer AB, Sweden) into one well of a flat bottom 96-well plate (#734-2327, VWR). To reduce contamination between cards, the puncher’s blade was cleaned with a H_2_O_dd_-wetted, and then a 70% EtOH-wetted, synthetic swab for every new row of the 96-well plate. The transferred discs were then subjected to 100 µl of PBST containing 1x PBS with 0.05% Tween20 and protease inhibitor cocktail (#04693116001, Roche). The discs were then incubated under gentle shaking (170 rpm) for 60 min at room temperature. The plates were then centrifuged for 3 min at 3000 rpm (Allegra X-12R, Beckman Coulter Inc.). A supernatant of 70 µl was transferred into a PCR plate (#732-4828, VWR) and sample eluates were stored at −20°C after the analysis. Protein concentrations of the eluates were determined using a Nanodrop spectrophotometer system (ND-1000, ThermoFisher) by measuring the absorbance at 280 nm in 2 µl per samples. The eluates were measured in triplicates and using the elution buffer as blank.

### Serology assays

#### Multiplexed serology

Proteins were covalently coupled to color-coded magnetic beads (MagPlex, Luminex Corp.) using either NHS/EDC coupling as described elsewhere *(25)*, or using an Activation Kit For Multiplex Microspheres (A-LMPAKMM-10RXN, SigmaAldrich) for proteins stored in Tris-based buffers. Anti-human IgG (309-005-082, Lot# 132463, Jackson ImmunoResearch), anti-human IgM (109-005-129, Lot#147777, Jackson ImmunoResearch), and anti-human IgA (GA-80A, Lot# 0017, Immune Systems Ltd) antibodies were coupled using NHS/EDC coupling and were used as controls in the assays. The beads were then mixed to create a suspension antigen bead array. Conjugation was confirmed using epitope-tag specific antibodies.

DBS eluates in 96-well plates were transferred to 384-well plates for the serological analysis. Eluates were diluted 1:2.5 in assay buffer containing 1x PBS with 0.05% Tween20 with 3% BSA (B2000-500, Lot# 08C5415, Saveen Werner) and 5% milk powder (70166-500G, Lot# BCBT8091, Sigma-Aldrich). Negative and positive control plasma samples were diluted in assay buffer 1:50 and 1:7.5 respectively. Per diluted sample, 35 µl were then incubated with 5 µl antigen bead array for 1 h at room temperature, shaking at 650 rpm, dark, followed by washing in 3×60 µl PBS-T 0.05% using an automated washing system (Biotek EL406). The beads were then resuspended in 50 µl detection buffer containing either anti-human IgG-R-PE (H10104, Lot# 2079224, Invitrogen) diluted to 0.4 µg/ml in PBS-T 0.05% or anti-human IgM-R-PE (#109-116-129, Lot# 137465, Jackson Immunoresearch) diluted 1:500 in PBS-T 0.05% diluted 1:500 in PBS-T 0.05%. The beads were then incubated for 30 min at room temperature under rotational shaking at 650 rpm in the dark. Prior to performing the readout on a FlexMap instrument (Luminex Corp) the plates were washed 3x 60 µl with PBS-T 0.05%, and 60 µl PBS-T were added into each well. The data was reported as median fluorescence intensity (MFI) values per antigen and sample. For each of the data points, at least 32 events per bead ID were collected.

#### ELISA

Seropositivity levels for human IgG were determined for S1 protein (EI 2606-9601 G, Lot# E200428BX, EuroImmun AG) and the N protein (EI 2606-9601-2 G, Lot# E200429BO, EuroImmun AG) according to the kit provider. Plasma samples were diluted 1:101, and DBS eluates were diluted 1:5 in the provided assay buffer.

### Affinity proteomics

Multiplexed protein analysis was performed at SciLifeLab’s Plasma Profiling facility in Stockholm using Olink panels Cardiovascular III (Product No 95611, Lot No B01116), Metabolism (Product # 95340, Lot # B01109), and Cardiometabolic (Product No 95360, Lot No B02504) according to manufacturer’s instructions (Olink Proteomcs AB). The analysis is based on the proximity extension assay technology *(26)*. In brief, 1 µl plasma sample were diluted 1:100, 1:20, or 1:2025 and as recommended while 1 µl DBS eluates were diluted 1:5, 1:1, or 1.101, respectively. These samples were then incubated with a pair of oligonucleotide-labeled antibodies for 92 analytes simultaneously. When an antibody pair binds its intended target, the oligonucleotides are brought in close proximity allowing for hybridization and DNA polymerization. These events procedure a reporter sequence to be quantified using a microfluidic real-time PCR instrument (Biomark HD, Fluidigm) to determine protein abundance as normalized protein expression (NPX) values.

### Data processing and statistical analyses

Data processing and analyses were performed in v.3.6.0 of R *(27)* or Julia *(28)*. For the SBA data of the study sets 1 and 2, MFI values were log transformed and normalized to adjust for background binding of human IgG as follows. A trend line was fitted to the bulk data which was assumed to represent the more frequent seronegative samples by regressing each protein profile against those reported for the internal negative control. This control was one population of beads subjected to the coupling procedure without the addition of any proteins, denoted as bare beads. To minimize the influence of the less frequent seropositive samples, we used a robust regression model with a Huber loss function *(29)* consisting of an L2 loss for errors smaller than 0.1 and L1 loss for larger errors, as well as an L2 penalty on regression parameters. This procedure was implemented in the Julia package MLJLinearModels.jl and applied to normalize the data on the basis of the assay background obtained from the bare bead with the peak of the seronegative samples centered at zero. This normalization converted raw MFI data to residuals from this regression and denoted as relative antibody titers.

#### Seroprevalence

The prevalence was estimated based on 3x SD and 6x SD added to the peak value of density plot applying Gaussian smoothing. The SD was the standard deviation of the values excluding 20% far from the density peak. For calculating the confidence interval (CI) of 95%, we assumed each set was a random sample from Stockholm population and applied normal approximation. Positive predictive value (PPV) was computed assuming the prevalence of the disease was 10%. When computing values for sensitivity, specificity and PPV, repeated measurement of positive/negative control samples were not taken into consideration. The UpSet plots were created by “UpSetR v. 1.4.0” R package.

#### Dimensionality reduction

PCA and UMAP analysis were performed using the R packages “stats v 3.6.0” and “umap v. 0.2.4.1” (https://CRAN.R-project.org/package=umap) respectively. For UMAP analysis a range of values for the hyperparameters “n_neighbors” and “min_dist” were tested with three repeats for each parameter combination. For each data set an UMAP layout representing the whole collection was chosen for visualization.

#### Affinity proteomics

The R package “limma v. 3.42.0” was used to determine the differentially abundant proteins from Olink’s protein analyses. NPX values were processed with the “lmFit” function. The reported p-values were corrected for multiple testing using the default setting “BH” (Benjamini-Hochberg) and reported FDR < 0.01 were deemed significant.

## Supplementary Materials

### Supplementary Text

- Assay Development
- Carry-over
- Precision
- Detectability

### Supplementary Figures

- Fig. S1. Detailed description of translational serology workflow and assays.
- Fig. S2. Carry-over assessment.
- Fig. S3. Limit of detection assessment.
- Fig. S4. Serology profiles from ELISA and multiplexed SBA assays in plasma and DBS.
- Fig. S5. Comparison of sample types and serology assays.
- Fig. S6. Participation in population-based home sampling.
- Fig. S7A. Multiplexed serology in home sampled blood from study set 1.
- Fig. S7B. Multiplexed serology in home sampled blood from study set 2.
- Fig. S8. IgG profiles of UMPA-selected seropositive and seronegative individuals.
- Fig. S9. IgM profiles of UMPA-selected seropositive and seronegative individuals.
- Fig. S9. Affinity proteomics analysis for differentially abundant proteins in DBS eluates of seropositive and seronegative individuals

### Supplementary Tables

- Table S1. Inter- and Intra-day variability.
- Table S2. Demographic characteristics of the feasibility study
- Table S3. Antigen-centric determination of IgG seropositivity.
- Table S4. Geographic selection for the population study.

### Supplementary Data File

- Index: Sheet Names and Read me
- Prevalence per Antigen (Set 1): List of antigens and seroprevalence performance (SBA assays)
- Prevalence per Antigen (Set 2): List of antigens and seroprevalence performance (SBA assays)
- Toptable_plasma_elisa : List of proteins and measured differences between seropositive and seronegative plasma samples from feasibility study (Olink assays)
- Toptable_olink_vDBS_vs_cDBS: List of proteins and measured differences between two types of DBS eluates from feasibility study (Olink assays)
- Toptable_olink_plasma_vs_DBS: List of proteins and measured differences between plasma and DBS eluates from feasibility study (Olink assays)
- Toptable_population_umap_groups: List of proteins and measured differences between seroposirive and seronegative plasma samples from population study (Olink assays)

## Data Availability

Normalized and anonymized serology data of the population studies will be deposited and can be made available for validation purposes and upon reasonable request to the corresponding authors.

## Acknowledgments

We thank all anonymous blood donors who volunteered to help us with this research project. We also thank A. Benckert, C. Stenfelt, A. Foley, T. Lundén, J. Lundén, E. Roxhed, V. Wennergren and X. Tian who volunteered to pack envelopes; Drs. C. Linder, A. Pohanka, S. Rosenborg from Karolinska hospital and Dr. S. Rautiainen-Lagerström at Danderyds hospital for assisting in obtaining samples. At SciLifeLab, we thank Dr. Anders Olsson, Dr. Maurice Michel, Kirill Mamonov und Florian Ortis for their continuous efforts on generating proteins and peptides. We thank the support from all members of the Plasma Profiling and Autoimmunity Profiling Facilities at SciLifeLab in Stockholm. Everyone at the Human Protein Atlas Project is acknowledged for their tremendous efforts.

## Author contributions

AB, MD, CM generated the immunoassay data and contributed to the experimental study design. LH, BiMe, SE, JA produced and provided viral antigens. SH, CT, and JD provided clinical samples. MC and BM developed the normalization approach. TDC, CET and MGH contributed to the experimental study design and analyzed the data. BeMu and GM provided expertise on virology and viral antigens. OB developed the sample preparation protocol. CF developed the sample preparation and assay protocol, supervised the lab work, and contributed to the experimental design. JMS and NR conceived and supervised the study. JMS wrote the manuscript with input and feedback from all authors. The final version of the manuscript was approved by all co-authors.

